# Infant Skincare in India: Clinical Practices, Perceptions, and Evolving Perspectives from a Healthcare Professional Survey

**DOI:** 10.64898/2026.09.08.26362227

**Authors:** Huma Rizwan, Bharat J. Parmar, Shruti Mukherjee, Shahani Begum, Pranita Ray, Prarthana Chatterjee

**Affiliations:** Clirnet Services Private Limited, West Bengal 700091, India; Zydus Medical College and Hospital, Gujarat 389151, India

**Keywords:** Infant Skincare, Atopic Dermatitis, Natural Skincare Product

## Abstract

**Objective:** Infant skin is highly susceptible to dermatitis, barrier dysfunction, and infections, necessitating safe and evidence-based skincare. This study assessed perspectives of Indian healthcare professionals (HCP) on infant skincare practices, product recommendations, prescribing behavior, and unmet needs in sensitive skin and atopic dermatitis management.

**Methods:** An online multicentric 4 independent cross-sectional survey was conducted involving 358 healthcare professionals across India, including pediatricians, dermatologists, and other clinicians, using structured questionnaires. The survey assessed prescribing behaviors, awareness of parents of the available products, and unmet needs in sensitive skin and atopic dermatitis management.

**Results:** The survey findings identified sensitive skin as the most frequently encountered infant skincare concern (91%), followed by eczema/atopic dermatitis (89%), and diaper rash (83%). Emollients were the most preferred first-line management strategy for mild-to-moderate sensitive infant skin (69%), followed by natural skincare products (63%), reflecting a strong preference for barrier-supportive and moisturizing therapies. Most HCPs preferred baby skincare products free from harsh chemicals (92.9%), while more than 90% routinely recommended natural/ayurvedic formulations. Additionally, Most HCPs were aware of cow ghee-based baby skincare products (93.8%) and considered barrier-focused early skincare important in preventing atopic dermatitis and the atopic march (81-87%), with 89% strongly endorsed consistent daily skincare as a key strategy for preventing flares and maintaining long-term control in infant atopic dermatitis.

**Conclusion:** Indian healthcare professionals favor preventive, barrier-focused infant skincare, with strong preference for natural and dermatologically safe formulations, while emphasizing the need for stronger clinical evidence in sensitive skin and atopic dermatitis management.

## Introduction

Infant skin differs structurally and functionally from adult skin and undergoes progressive maturation during the early years of life. The epidermal barrier in neonates and infants is thinner, exhibits higher transepidermal water loss, reduced hydration, making it more vulnerable to irritation, dryness, and inflammatory dermatoses. ^[1⍰2]^ Consequently, dermatological concerns such as sensitive skin, diaper dermatitis, eczema, allergic reactions, and atopic dermatitis are frequently encountered during infancy and represent a substantial clinical burden for both healthcare professionals and parents. Atopic dermatitis affects 20% to 25% of children and commonly begins within the first year of life, often impairing sleep, comfort, and quality of life. ^[3,5]^

Increasing understanding of skin barrier dysfunction and cutaneous inflammation has shifted infant skincare practices from a predominantly reactive approach toward preventive and barrier-focused strategies. Current recommendations emphasize gentle cleansing, regular moisturization, and maintenance of epidermal barrier integrity as key components of infant skincare, particularly in children with sensitive skin and atopic tendency. ^[6]^ Consistent use of emollients and barrier-supportive skincare has been associated with improved skin hydration, reduced flare frequency, and enhanced tolerability. Early-life skincare is also increasingly recognized for its potential role in influencing the progression of the atopic march through preservation of skin barrier function and reduction of allergen penetration. ^[6-8]^

Alongside evolving clinical practices, parental awareness regarding ingredient safety and product tolerability has substantially increased. Parents increasingly prefer products perceived as natural, gentle, dermatologically tested, and free from harsh chemicals. This growing demand has contributed to increased interest in plant-based and traditionally derived skincare ingredients. In India, traditional infant skincare practices have long included the use of natural ingredients such as cow-derived ghee, which has historically been valued for its emollient, soothing, and protective properties. Modern infant skincare formulations increasingly attempt to integrate such traditional ingredients with contemporary standards of dermatological safety, tolerability, and clinical validation. ^[9,10]^

Despite the availability of numerous baby skincare products, variability persists in clinical practice regarding product recommendations, prescribing behavior, evidence expectations, and perceptions of natural formulations. Healthcare professionals play a central role in guiding parental decisions regarding infant skincare, especially in vulnerable populations with sensitive skin or atopic dermatitis. Therefore, understanding clinicians’ experiences, prescribing preferences, confidence in preventive skincare, and expectations regarding product safety and efficacy is essential for aligning skincare practices with evidence-based dermatologic care. ^[6,11]^

Additionally, while natural and plant-based formulations are increasingly favored in routine practice, clinicians continue to emphasize the importance of scientific evidence, dermatological testing, and real-world clinical outcomes before recommending such products. Insights into clinician satisfaction and unmet therapeutic needs may further support the development of safer and more effective infant skincare strategies. ^[2,11,12]^

The present four independent cross-sectional surveys study was conducted to evaluate evolving infant skincare practices among healthcare professionals in India, with particular focus on sensitive skin and atopic dermatitis management, preventive barrier-focused skincare, product recommendation behavior, and perceptions toward natural formulations.

## Methods

### Study Design and Participants

This was a multicentric, cross-sectional descriptive survey conducted among healthcare professionals involved in the care of newborns and infants across India. A total of 358 practitioners participated in the survey, predominantly pediatricians, along with representation from dermatology, gynecology, and general practice. Participants were recruited from diverse geographic regions, including Tamil Nadu, Maharashtra, Gujarat, Delhi, Odisha, Telangana, Karnataka, Uttar Pradesh, Madhya Pradesh, West Bengal, and other states across India. Data were collected using structured questionnaires administered online. To maintain participant confidentiality, all personally identifiable information, including names and addresses, was excluded from the analysis. Consequently, no individual healthcare professional is identifiable in this report.

### Survey Overview

**Table 1:**
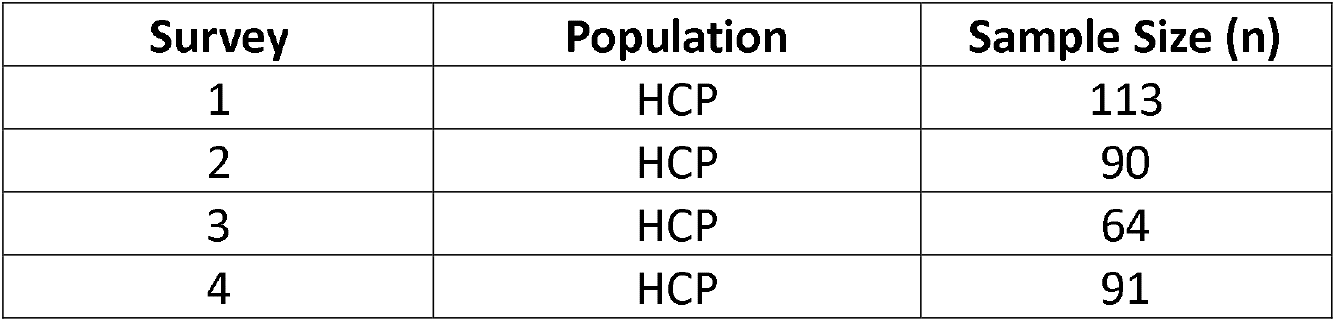
Outline of survey populations and sample sizes.

| Survey | Population | Sample Size (n) |
| --- | --- | --- |
| 1 | HCP | 113 |
| 2 | HCP | 90 |
| 3 | HCP | 64 |
| 4 | HCP | 91 |

### Data Handling and Analysis

Data were analyzed using descriptive statistics. Categorical variables were summarized as frequencies and percentages. For multiple-response questions, each selected response was analyzed independently and expressed as a percentage of the total study population.

## Results

### Prevalence and Management

Most HCPs (91%) reported that sensitive skin is a very common concern in infants with 25–50% of infant patients present with skin-related concerns. Eczema/atopic dermatitis (89%) and diaper rash (83%) were the most frequently encountered conditions [Figure 1], while emollients (69%) and natural skincare products (63%) were the preferred first-line management strategies for mild-to-moderate sensitive infant skin [Figure 2]. A majority of clinicians (89%) strongly endorsed consistent daily skincare for preventing flares in infant sensitive skin and atopic dermatitis, 81% believed early-life skincare strongly influences the risk and progression of atopic dermatitis, while 73% preferred initiating preventive skincare from birth in high-risk infants [Figure 3]. Additionally, 87% strongly supported barrier-focused skincare as a preventive strategy against the atopic march, reflecting high clinical confidence in its long-term protective role.

**Figure 1:**
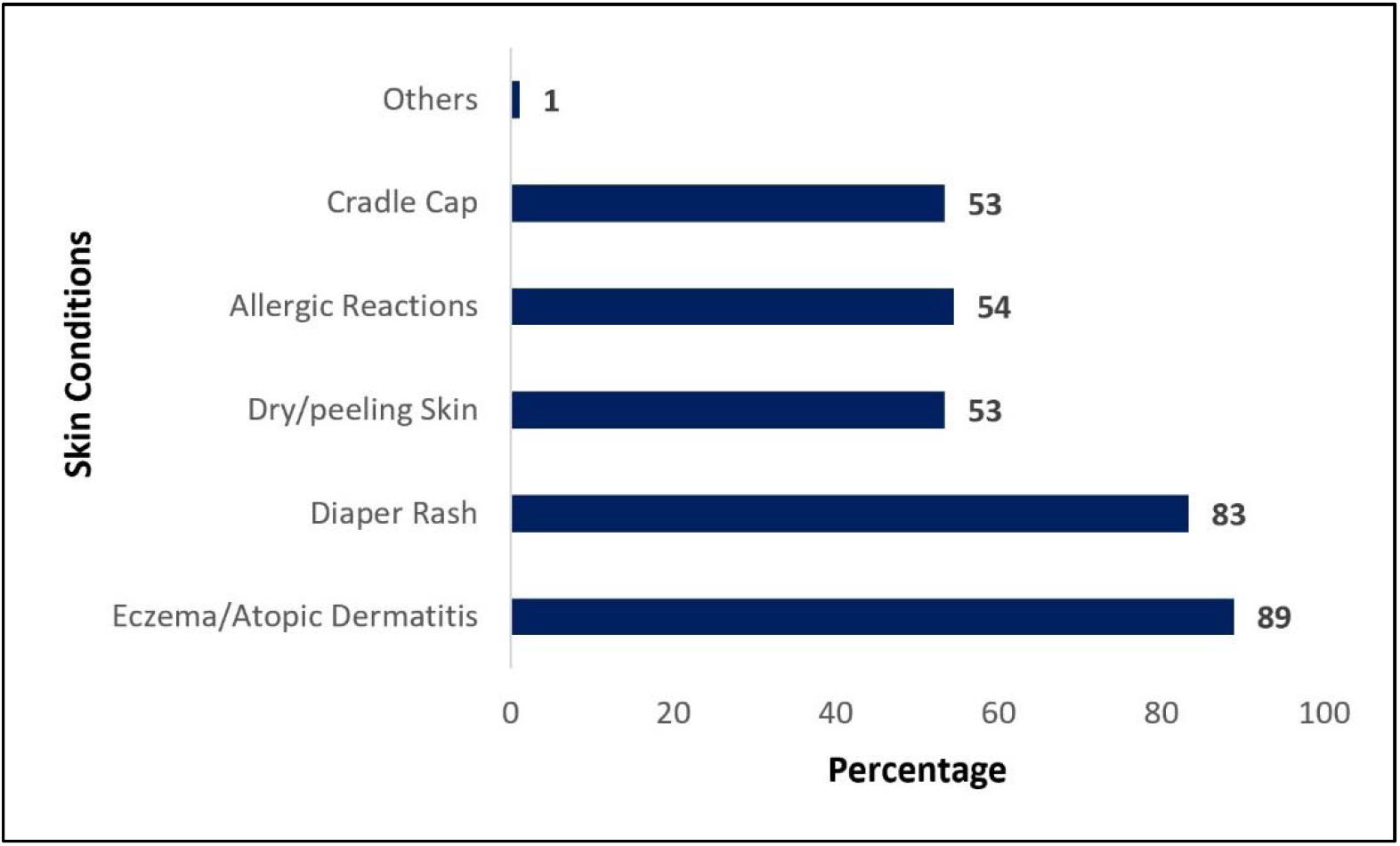
Common skin condition in infants. The figure summarizes HCP-reported responses regarding the frequency of various skin conditions observed in their practice. Eczema/atopic dermatitis and diaper rash represent the vast majority of reported cases. Responses are presented as percentages for each category. (Percentages sum to >100% due to multiple-choice response options).

**Figure 2:**
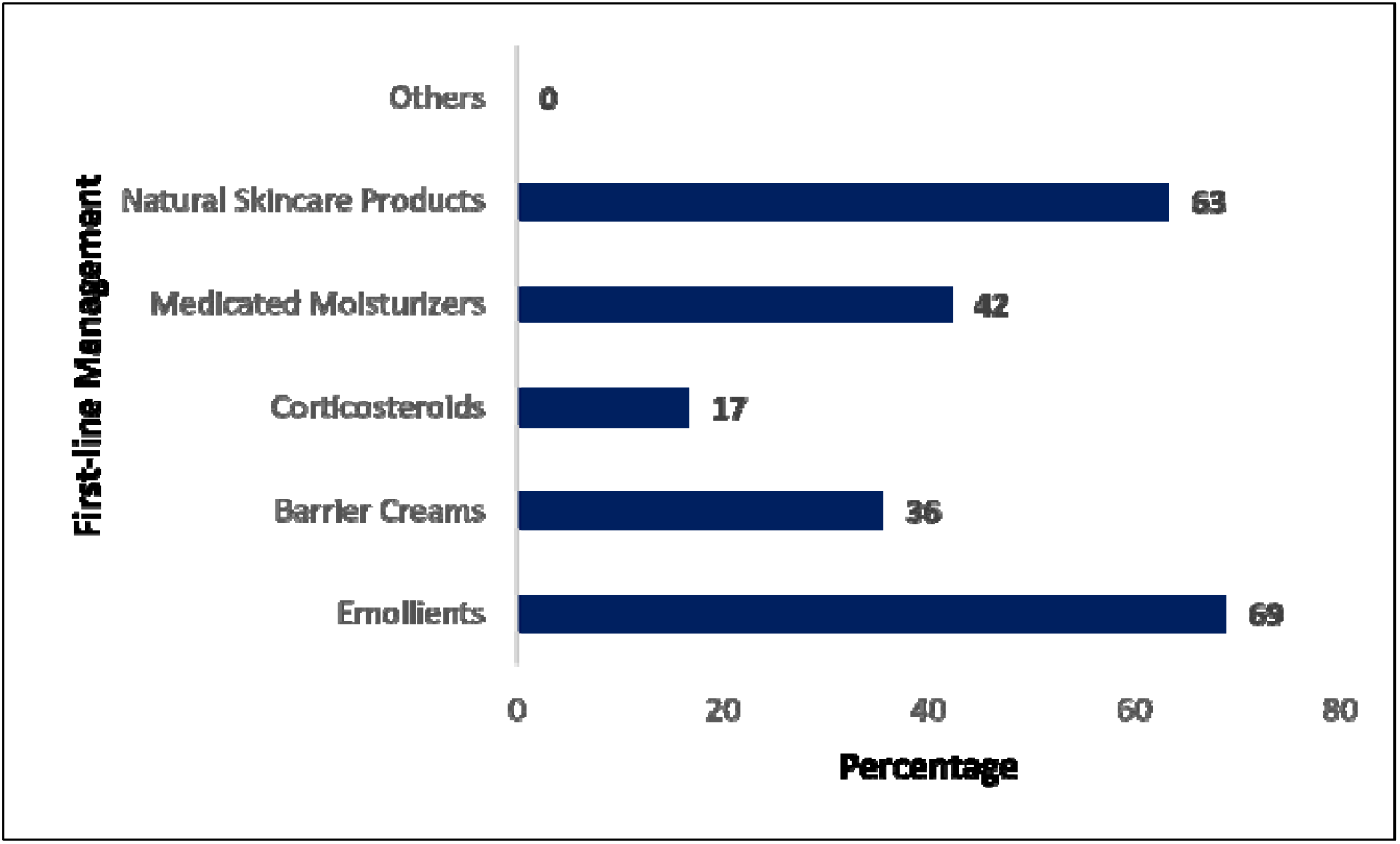
Primary infant skincare management. The figure summarizes HCP-reported responses regarding their preferred first-line management strategies. Emollients and natural skincare products emerged as the top two first-line recommendations. Responses are presented as percentages for each category. (Percentages sum to >100% due to multiple-choice response options).

**Figure 3:**
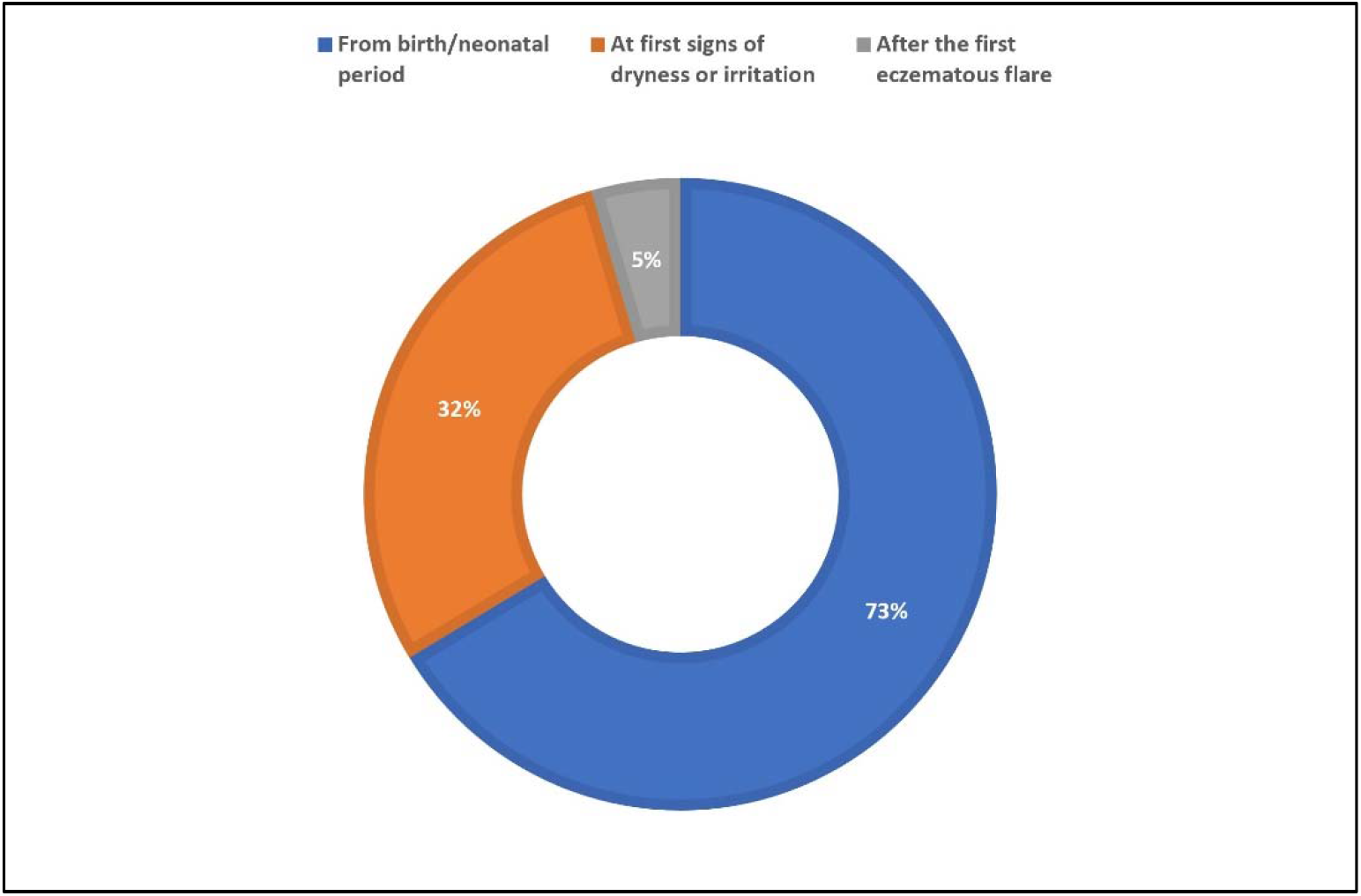
Initiation of preventive infant skincare. The figure summarizes HCP-reported responses regarding the optimal time to introduce skincare routines in infants. Initiation from birth/neonatal period emerged as the leading clinical recommendation. Responses are presented as percentages for each category. (Percentages sum to >100% due to multiple-choice response options).

### Product Preferences and Prescribing Factors

When asked which attributes are essential in a product for babies with sensitive skin and newborns, most HCPs preferred baby skincare products free from harsh chemicals (92.9%), dermatologically tested (69%), and enriched with natural ingredients (65.5%) [Figure 4].

**Figure 4:**
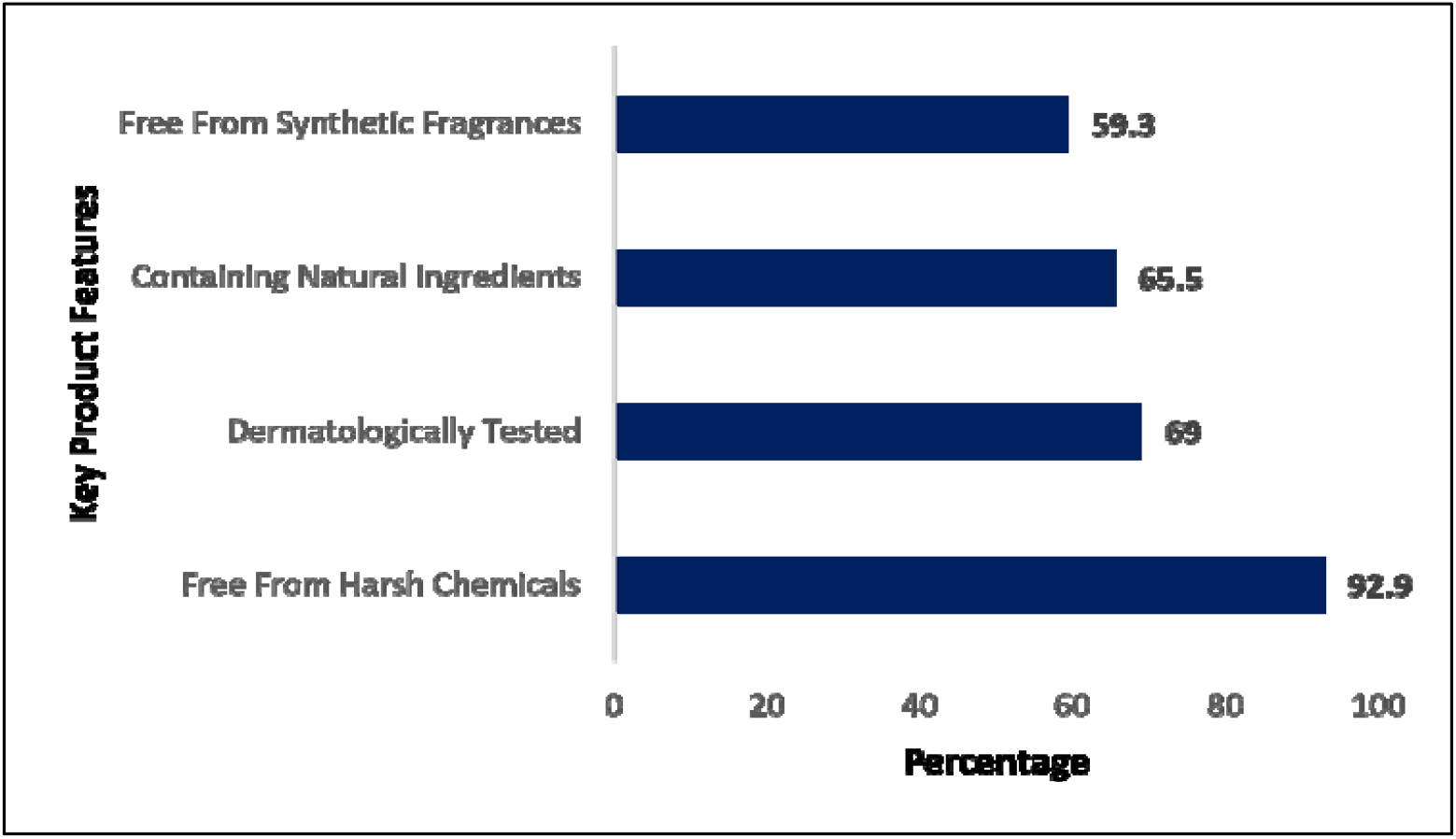
Preferred attributes of baby skincare product. The figure summarizes HCP-reported responses regarding the most important factors influencing their product recommendations. Safety and formulation purity, led by the absence of harsh chemicals, were the top considerations for healthcare providers. Responses are presented as percentages for each category. (Percentages sum to >100% due to multiple-choice response options).

### Product Recommendation Criteria

Clinical efficacy (94%), ingredient safety (76%), hypoallergenic claims (72%), and brand trust (68%) were the key factors influencing baby skincare product recommendations [Figure 5]. Most clinicians (86%) preferred products specifically formulated for sensitive skin, while 73% were comfortable recommending clinically safe, plant-based formulations.

**Figure 5:**
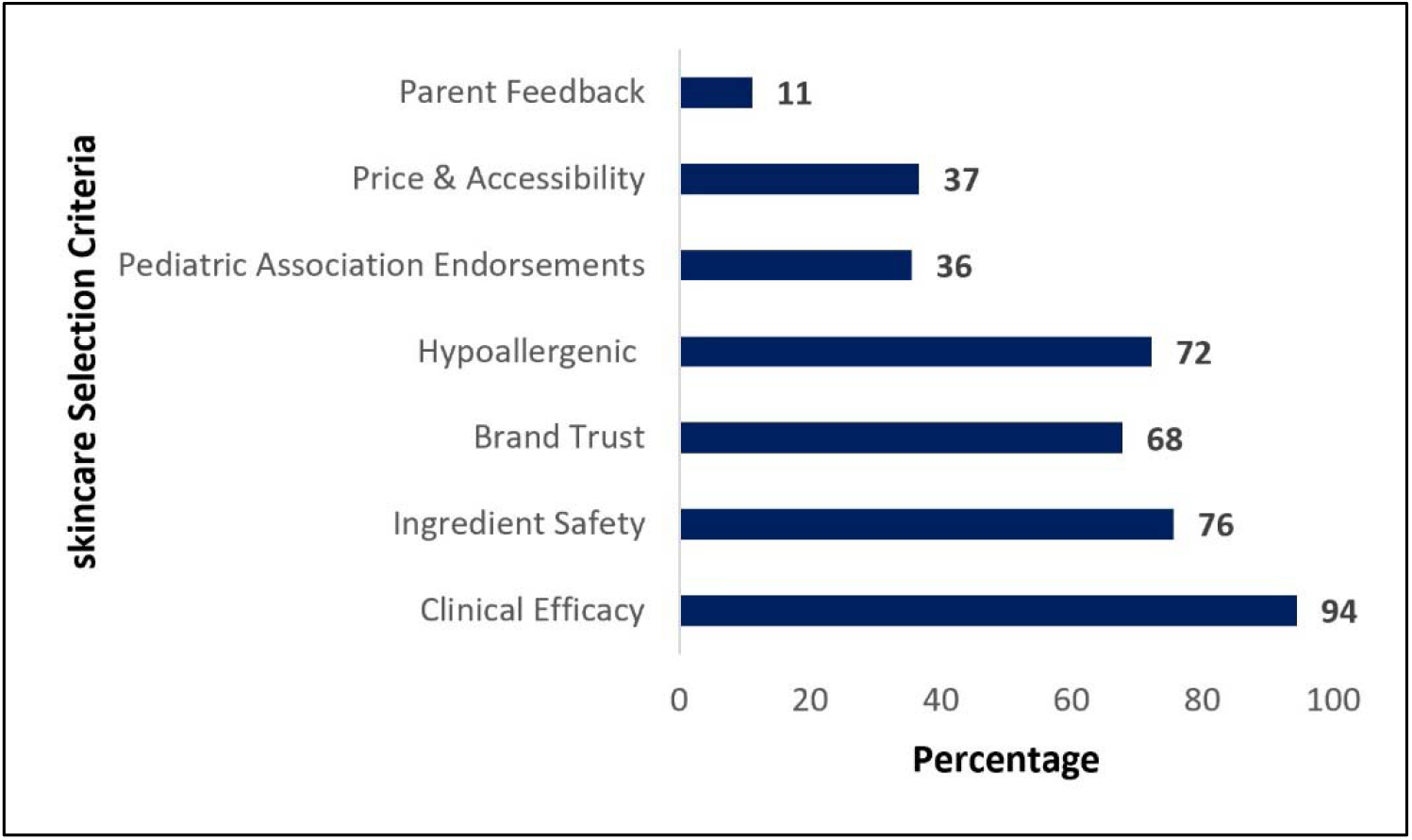
Factors influencing baby skincare product recommendations. The figure summarizes HCP-reported responses regarding the key factors guiding their product selection. Clinical efficacy and ingredient safety emerged as the predominant clinical drivers. Responses are presented as percentages for each category. (Percentages sum to >100% due to multiple-choice response options).

### Preventive Natural Skincare and Perception of Cow-Ghee–Based Products

Most clinicians (80%) used natural skincare products as preventive and maintenance strategies to reduce flare occurrence in infants, with safety profile and tolerability (75%) being the leading factors influencing recommendation confidence. Awareness of pure cow-ghee–based baby skincare products was high (93.8%), and clinicians commonly perceived benefits such as anti-inflammatory effects (31.9%), moisturization (31%), soothing of irritation and rash (15%), and support of skin barrier protection [Figure 6].

**Figure 6:**
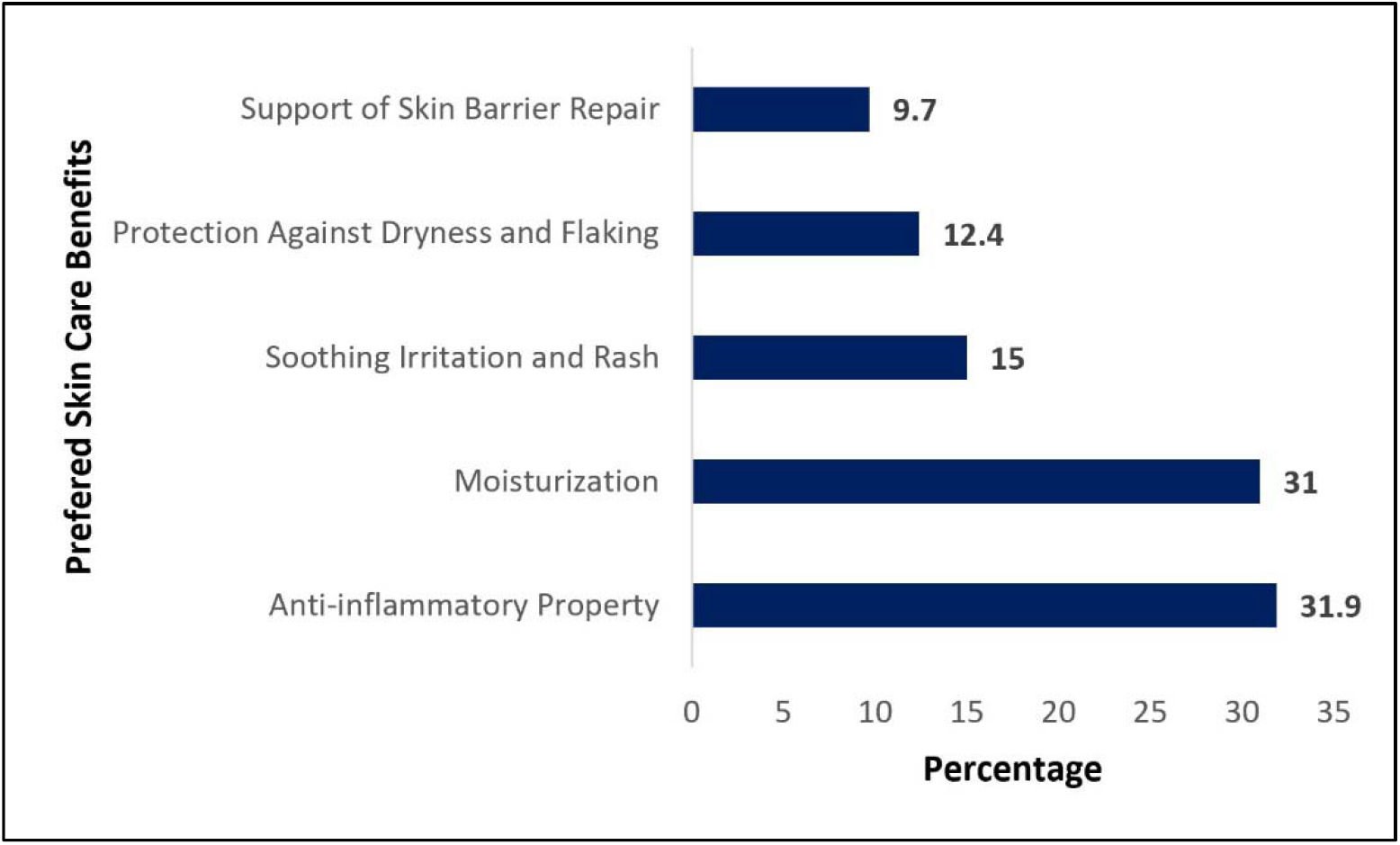
Benefits of cow-ghee-based product. The figure summarizes HCP-reported responses regarding the single most important clinical outcome expected from their recommended skincare interventions. Anti-inflammatory action and moisturization of ghee emerged as the two primary outcomes anticipated by healthcare providers. Responses are presented as percentages for each category. (Percentages sum to >100% due to multiple-choice response options).

### Satisfaction with Current Options & Unmet Needs

A majority of clinicians (84%) reported satisfaction with currently available skincare options for infants with sensitive skin and atopic tendency. However, longer-lasting moisturization (75%), improved flare prevention (25%), and better tolerability on inflamed skin (19%) were identified as key unmet clinical needs [Figure 7].

**Figure 7:**
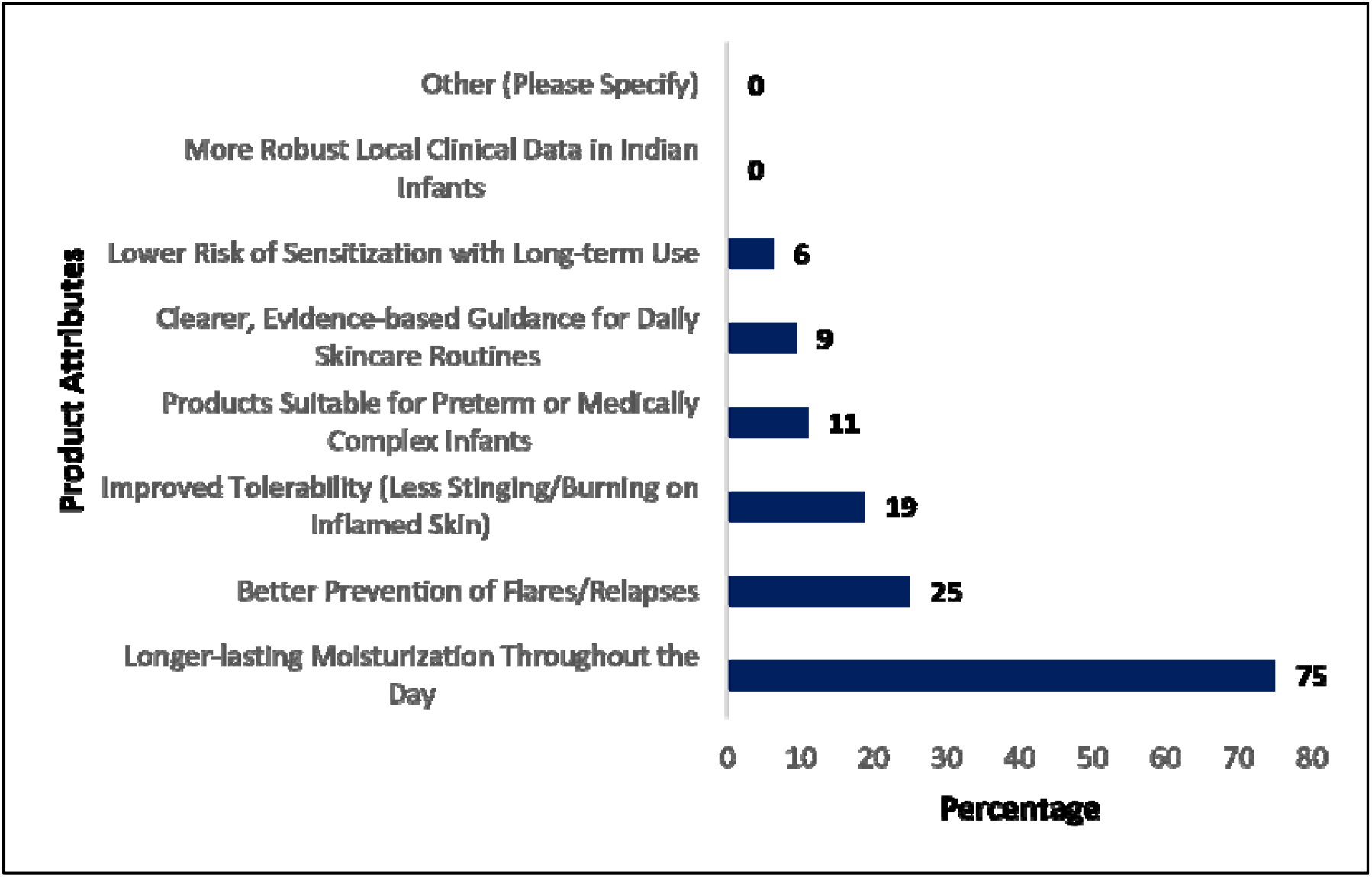
Unmet needs for infant skincare products. Among clinicians, longer-lasting moisturization (75%), improved prevention of flares/relapses (25%), and better tolerability on inflamed skin (19%) were the most commonly reported unmet needs for daily skincare routines, and reduced sensitization risk with long-term use.

## Discussion

Infant skin is highly susceptible to irritation, dryness, and barrier dysfunction because of its structural and functional immaturity. Appropriate skincare during early life therefore plays an important role in maintaining epidermal integrity and preventing inflammatory skin conditions. The present survey provides insights into evolving infant skincare practices among Indian healthcare professionals, particularly regarding sensitive skin management, preventive skincare strategies, and the growing preference for natural and barrier-supportive formulations. ^[13]^

In the current study, majority of healthcare professionals considered sensitive skin a common concern in infants, with eczema/atopic dermatitis and diaper rash being the most frequently encountered conditions. These findings are consistent with previous reports showing that inflammatory and barrier-related dermatoses are highly prevalent during infancy and early childhood. ^[3,4]^ The increased vulnerability of infant skin is attributed to higher trans-epidermal water loss, thinner stratum corneum, and incomplete barrier maturation, making preventive skincare particularly important during the neonatal period. ^[1,2]^

Most respondents preferred emollients and natural skincare products as first-line approaches for mild-to-moderate sensitive skin. This observation aligns with current recommendations emphasizing regular moisturization and barrier repair as essential components of infant skincare and atopic dermatitis management. ^[14]^ Consistent use of moisturizers has been shown to improve skin hydration, strengthen barrier function, and reduce flare frequency in infants predisposed to eczema. ^[8]^ In the present survey, clinicians also strongly supported daily skincare regimens involving gentle cleansing and regular moisturization, further reinforcing the role of preventive barrier care in routine pediatric practice. An important finding of this study was the strong clinician preference for skincare products that are free from harsh chemicals, dermatologically tested, and enriched with natural ingredients. Healthcare professionals also considered clinical efficacy, ingredient safety, hypoallergenic claims, and brand trust as major determinants influencing product recommendations. These findings suggest that clinicians increasingly favor evidence-based and safety-oriented formulations for infant skincare.

Natural and traditionally derived formulations are gaining popularity in infant skincare because of their perceived safety and tolerability. In the present study, most respondents were aware of cow-ghee–based baby skincare products and considered them safe for infants with sensitive skin. Clinicians commonly associated these products with moisturization, anti-inflammatory activity, soothing effects, and barrier support. Traditional Indian skincare practices have long incorporated cow-derived ghee for neonatal massage and moisturization because of its emollient and protective properties. ^[10]^ The growing integration of such traditional ingredients into modern dermatologically evaluated formulations reflects the increasing convergence of traditional practices with evidence-based skincare approaches.

The survey also highlighted increasing emphasis on preventive dermatology and early barrier-focused intervention. Most respondents believed that early-life skincare significantly influences the risk and progression of atopic dermatitis and supported initiation of preventive skincare from birth in high-risk infants. Emerging evidence suggests that maintenance of skin barrier integrity during infancy may reduce inflammatory responses associated with the atopic dermatitis. ^[8^⍰^15]^ The strong support for barrier-focused skincare observed in this study indicates growing clinician awareness regarding the long-term benefits of early preventive intervention.

Although overall satisfaction with currently available infant skincare products was high, clinicians identified several unmet needs, including long-lasting moisturization, improved flare prevention, and enhanced tolerability in inflamed skin. These findings indicate the need for continued innovation in infant skincare formulations with improved barrier-supportive properties, sustained hydration, and robust clinical validation. The study has certain limitations. Being a questionnaire-based survey, the findings are dependent on clinician perceptions and self-reported practices, which may introduce reporting bias. Additionally, the cross-sectional design limits assessment of longitudinal clinical outcomes. Nevertheless, the study provides valuable real-world insights into current infant skincare trends and prescribing practices among healthcare professionals across India.

Overall, the findings demonstrate a clear shift toward preventive, barrier-focused, and natural skincare approaches in infant dermatology, with increasing emphasis on safety, tolerability, and evidence-based product selection for sensitive skin and atopic dermatitis management.

## Conclusion

Clinicians in India are increasingly integrating natural, barrier-focused products, prioritizing “free-from” formulations and traditional ingredients like cow ghee into routine infant care. While overall satisfaction with current products is high, persistent unmet needs for durable moisturization and improved flare prevention remain. To ensure long-term adoption, manufacturers must prioritize clinical trials and transparent ingredient lists. Enhancing locally relevant clinical evidence will be crucial for optimizing infant skincare and modifying disease progression.

## Data Availability

All data produced in the present work are contained in the manuscript

